# Presentation and Types of Childhood Cancer at the Muhimbili National Hospital (MNH), 2023

**DOI:** 10.1101/2025.01.28.25321242

**Authors:** Malaz Adam Ali Salih Arga, Rehema H. Laiti, Ghada Omer Hamad Abd El Raheem, Yasar Hammor

## Abstract

**BACKGROUND:** In sub-Saharan Africa in Tanzania, with an incidence of 1.4/100,000 cases. A lot of cancer cases in Tanzania are not diagnosed until later because there aren’t many healthcare institutions offering cancer-related care and treatment. Cancer that manifests in its late stages not only reduces survival chances but also places a heavy load on the healthcare system. a lower-middle-income nation, has numerous barriers to receiving treatment for children’s malignancies. The research aims to study the clinical presentation and types of childhood cancer at the Muhimbili National Hospital.

**METHODS:** A hospital-based cross-sectional survey and convenience sample technique were used to assess the presentation and association factors of childhood cancer. Descriptive statistics and statistical analysis to assess associations among variables were performed through chi2 and one-way ANOVA. A p-value of <0.05 was considered significant.

**RESULTS:** 141 patients were assessed; the most common types of cancer were Blastoma and Leukaemia between 1 and 5 years. The Sukuma tribe had the highest percentage of patients. Most of the patients came from Dar es Salaam. The majority of patients presented with masses and abdominal distention. There was a statistically significant difference between family history and the type of cancer, between the stage of disease and response to treatment, and between the ages across types of cancer. There is no statistical difference or association between the type of cancer and the history of chronic illness or exposure.

**CONCLUSION:** Childhood cancer has high mortality and morbidity in Tanzania. Most of the patients die before 5 years old, and patients come to the hospital with a late-stage disease with serious symptoms. Other patients didn’t complete treatment, which may be due to financial problems or a low level of education among carers.

## INTRODUCTION

Cancer is the most common disease-related cause of death for children. Childhood malignancies, which occur in the setting of tissues that are actively developing, are disorders of dysregulated development. It is the second-leading cause of death among children in the United States and other developed countries, after accidents. Pediatric cancers include Leukemia, Lymphoma, Central Nervous System tumors, bone and soft tissue Sarcomas, Neuroblastoma, Retinoblastoma, Rhabdoid tumors, Liver tumors, Kidney tumors, Germ cell tumors, and other rare cancers. [1]

The International Childhood Cancer Cohort Consortium (I4C) was conducted to examine the relationship between widespread environmental exposures and childhood leukemia. All cohort studies participating in I4C have or will include various exposure measurements in their protocols. These include: parental health interventions (e.g., infectious diseases); occupational, household, and lifestyle exposures (e.g., smoking, drug use, diet); child health measures (e.g., growth and infection); housing; and lifestyle exposures (e.g., dietary or chemical exposure). Exposure to pesticides is one of several environmental risk factors thought to increase the likelihood of developing paediatric cancer. The International Agency for Research on Cancer (IARC) has designated a few chemicals as human carcinogens. Early infancy and the perinatal period are crucial windows of increased vulnerability to environmental exposures. Before conception, during pregnancy, and after birth, a child may be impacted by a parent’s professional exposure to pesticides. [2, 17]

At Tanzania’s largest paediatric oncology centre, Muhimbili National Hospital (MNH), paediatric sarcomas constitute a significant subset of childhood mors. Treatment guidelines that have been updated based on medication availability and surgical and radiation therapy resources are used to guide management. These protocols are derived from clinical trials conducted by the International Society of Pediatric Oncology and the Children’s Oncology Group. There aren’t any baseline statistics available on Tanzania’s paediatric sarcoma frequency or kinds at this time. To better target treatment procedures and resources towards the more prevalent paediatric sarcomas in Tanzania. [18]

In sub-Saharan Africa, Tanzania, a lower-middle-income nation, has numerous barriers to receiving treatment for children’s malignancies. In the country, there are only six paediatric oncologists: three of them work in Dar es Salaam, the capital city, and one at the Kilimanjaro Christian Medical Centre (KCMC), in the northern region. Moreover, for every 1.8 million people, there is only one pathologist. In comparison, there is one pathologist and almost 2,000 pediatric oncologists per 20,638 residents in the United States. [19]

Nearly 80% of children with cancer reside in low- and middle-income countries (LMICs), where access to treatment is frequently limited or prohibitively expensive. Because of this, only between 15 and 45% of these kids end up alive, while over 80% of kids in high-income nations do. Children in low- and middle-income countries have a low treatment rate and high mortality, making palliative care less likely to be important in paediatric oncology because paediatric oncologists are not aware of palliative care and the number of services has decreased. Providing palliative care. [23, 24]

## METHODOLOGY

Hospital-based cross-sectional retrospective data and an observational study were used to assess the presentation and association factors of childhood cancer. The study was carried out in the paediatric oncology unit of Muhimbili National Hospital Population: Children under 18 years of age were diagnosed with cancer or haematologic malignancies in the paediatric oncology department of the Muhimbili National Hospital (MNH). Inclusion criteria: children with histological or cytological diagnosis of cancer or haematologic malignancies from birth until 18 years old in Muhimbili National Hospital (MNH). The sample size was got from the admission book. of year 2023, it is the most accurate resource for all admission patients in paediatric oncology department in Muhimbili National Hospital. By used Convenience sampling technique to determine the sample size. The sample size was total coverage within four months in the middle of the year 2023 (May, June, August, and July), the sample size was 141 patients.

I got approval from the University of Medical Science and Technology on 11/12/2023, and I sent it to Muhimbili National Hospital. After that, I started the data collection from 18/12/2023 to 31/3/2024, collecting data from the Muhimbili National Hospital Paediatric Oncology Department in Dar es Salaam, Tanzania.

Data was collected from the patient’s files. Use the data collection tool attached below. I entered the data; quality control, analysis procedures, and computerised packages were used. The data was checked and validated for analysis. The variables were demonstrated numerically (mean, standard deviation, and median) and graphically (graphs, charts, and frequency tables for estimating proportions). The data collected was analysed using Microsoft Excel and the statistical package for social science (SPSS 23), and we used Geographic Information System (GIS); ArcGIS 10.3 was used to develop the map at the level of the Tanzania region to do a disruption map of the patients. The result was discussed and compared with correlated studies. The association between variables was determined through chi-square tests, and an analysis of variance (ANOVA) was used to determine the association between categorical variables and numerical variables. All Statistical tests will be considered significant when the p-value is < 0.05.

## RESULTS

A total of 141 pediatric cancer patients aged 0-18 years were analyzed in this cross-sectional retrospective study at Muhimbili National Hospital over four months. Most patients were between 1 and 5 years old (mean age 2.59 years, SD 0.942). There was a male predominance (n=80, 56.7%). The majority of caregivers had primary school education (n=51, 50%). Most patients came from the Dar es Salaam region (n=43, 30.5%) and the Pwani region (n=13, 9.2%). The most common tribes were Sukuma (n=14, 10.3%), Chagga (n=8, 5.9%), Makonde (n=8, 5.9%), and Waha (n=7, 5.1%).

The most common cancer types were Blastoma (n=41, 29.1%), Leukemia (n=37, 26.2%), and Sarcoma (n=24, 17%). In 64 patients with known cancer stage, most were in Stage 3 (n=32, 50%) or Stage 4 (n=21, 32.8%). Over half of the patients did not present with metastatic cancer (n=81, 57.4%), while the rest did (n=58, 41.1%), with the lung (n=31, 43.7%) and central nervous system (n=14, 19.7%) being the most common sites. Common presenting symptoms were swelling (n=47, 33.8%), abdominal distention (n=33, 23.7%), pain (n=29, 20.9%), and fever (n=24, 17.3%).

Most patients (n=113, 80.1%) were referred from other hospitals, mainly Kilimanjaro Christian Medical Centre (n=9, 6.4%) and Mnazi Mmoja Hospital (n=8, 5.7%). New admissions accounted for 19.9% (n=28). The most common cancer subtypes were Wilm’s tumor (n=18, 12.8%), Retinoblastoma (n=13, 9.2%), and Acute Myelogenous Leukemia (n=10, 7.1%).

Over half of the patients did not have a history of chronic illness (n=126, 89.4%). The most common chronic illnesses were hypertension (n=6, 4.3%) and sickle cell disease (n=4, 2.8%). Most patients did not have a family history of cancer (n=130, 95.6%) or exposure history (n=124, 87.9%). Tuberculosis was the most common infectious exposure (n=9, 6.4%).

Chemotherapy was the most common treatment (n=72, 51.1%), followed by chemotherapy with surgery (n=28, 19.9%). However, the majority of patients died (n=36, 25.5%) or did not complete treatment (n=30, 21.3%).

Cross-tabulation analyses revealed no significant association between gender, tribe, or patient admission with cancer type. There was no significant difference in metastasis based on caregiver education level. However, there was a significant association between family history and cancer type (p=0.004).

Age was significantly associated with cancer type (p=0.000), with Blastoma and Leukemia being most common in children aged 1-5 years. There was no significant difference in treatment response based on age.

An independent t-test showed a significant difference between disease stage and treatment response (p=0.006), but no significant difference between disease stage and caregiver education level.

**Table 4.1:** Demographic characteristics of the patients.

| **Gender** | Frequency | % |
| --- | --- | --- |
| Male | 80 | 56.7 |
| female | 61 | 43.3 |
| **Age** |  |  |
| 6-10 years | 33 | 23.4 |
| >10 years | 29 | 20.6 |
| <1 years | 12 | 8.5 |
| Mean age | 2.59 |  |
| **Education status of the caregiver** |  |  |
| primary school | 51 | 50.0 |
| secondary school | 19 | 18.6 |
| illiterate | 16 | 15.7 |
| college\university | 15 | 14.7 |
| Other | 1 | 1.0 |
| Total | 102 | 100.0 |
| Missing | 39 | 27.7 |
| **Tribe** |  |  |
| Sukuma | 14 | 10.3 |
| Chagga | 8 | 5.9 |
| Makonde | 8 | 5.9 |
| Waha | 7 | 5.1 |
| Gogo | 6 | 4.4 |
| Masai | 6 | 4.4 |
| Others | 87 | 60.3 |
| Missing | 5 | 3.7 |
| Total | 141 |  |

**Table 4.2:** Cancer Characteristics.

| **Type of cancer** | **Frequency** | **%** |
| --- | --- | --- |
| Blastoma | 41 | 29.1 |
| Leukemia | 37 | 26.2 |
| Sarcoma | 24 | 17.0 |
| Lymphoma | 19 | 13.5 |
| Carcinoma | 5 | 3.5 |
| Teratoma | 5 | 3.5 |
| Astrocytoma | 4 | 2.8 |
| Hemangioma | 2 | 1.4 |
| Anterior mediastinal mass | 1 | .7 |
| Blastoma. Sarcoma | 1 | .7 |
| Cystic Hygroma | 1 | .7 |
| Multiple Endocrine Neoplasia | 1 | .7 |
| **Stage** |  |  |
| 1 | 8 | 12.5 |
| 2 | 3 | 4.7 |
| 3 | 32 | 50.0 |
| 4 | 21 | 32.8 |
| Missing | 77 | 54.6 |
| **Metastasis** |  |  |
| No | 81 | 57.4 |
| Yes: | 58 | 41.1 |
| Abdomen | 1 | 1.4 |
| Bone | 4 | 5.6 |
| Bone mar | 3 | 4.2 |
| Central Nervous system | 14 | 19.7 |
| Lymphadenopathy | 2 | 2.8 |
| Liver | 9 | 12.7 |
| Lung | 31 | 43.7 |
| Tumour lysis syndrome | 1 | 1.4 |
| Missing | 2 | 2.8 |
| **Presentations** |  |  |
| Swelling | 47 | 33.8 |
| Abdominal Distention | 33 | 23.7 |
| Pain | 29 | 20.9 |
| Fever | 24 | 17.3 |
| Generalize Body Weakness | 17 | 12.2 |
| Bleeding tendency | 10 | 7.2 |
| Whitish pupillary reflex | 9 | 6.5 |
| Headache | 9 | 6.5 |
| Vomiting | 7 | 5 |
| Protrusion of the eye | 6 | 4.3 |
| Cough | 6 | 4.3 |
| Missing | 2 | 1.4 |

**Table 4.3:** patient admission at hospital.

|  | Frequency | % |
| --- | --- | --- |
| New admission | 28 | 19.9 |
| Kalamnjaro christian medical centre | 9 | 6.4 |
| Mnazi Mmoja Hospital | 8 | 5.7 |
| Temeke Regional Hospital | 7 | 5.0 |
| Amana Regional Referral hospital | 6 | 4.3 |
| Benjamin Mkapa Hospital | 6 | 4.3 |
| Bugando Hospital | 5 | 3.5 |
| Comperhensive Community Based Rehabilitation in tanzania Hospital | 5 | 3.5 |
| Muhimbili orthopedic institute | 5 | 3.5 |
| St.Francis Referral Hospital | 4 | 2.8 |
| Mbeya Zonal Referral Hospital | 3 | 2.1 |
| Morogoro Regional Referral Hospital | 3 | 2.1 |
| St.Benedict Ndanda Referral Hospital | 3 | 2.1 |
| St.Gaspar Referral and Teaching Hospital | 3 | 2.1 |
| Missing | 1 | 0.7 |
| Total | 141 |  |

**Table4.4:** Subtype of the cancer.

|  | Frequency | % |
| --- | --- | --- |
| Wilm's tumor | 18 | 12.8 |
| Retinoblastoma | 13 | 9.2 |
| Acute Myelogenous Leukemia | 10 | 7.1 |
| Osteosarcoma | 9 | 6.4 |
| Acute Lymphocytic Leukemia (B-Cell) | 8 | 5.7 |
| Acute promyelocytic Leukemia | 6 | 4.3 |
| Hodgkin Lymphoma | 6 | 4.3 |
| Neuroblastoma | 5 | 3.5 |
| Classical Hodgkin Lymphoma | 4 | 2.8 |
| Acute Lymphocytic Leukemia | 3 | 2.1 |
| Burkitt's Lymphoma | 3 | 2.1 |
| Rhabdomyosarcoma (Embryonal) | 3 | 2.1 |
| Sacroccygeal Teratoma | 3 | 2.1 |
| Soft tissue sarcoma | 3 | 2.1 |
| Acute Lymphocytic Leukemia. Acute Myelogenous Leukemia | 2 | 1.4 |
| Acute Myelogenous Leukemia (AML) | 2 | 1.4 |
| Anaplastic Large cell Lymphoma | 2 | 1.4 |
| Astrocytoma (Glioma) | 2 | 1.4 |
| Astrocytoma (polycystic) | 2 | 1.4 |
| Chronic Myelogenous Leukemia | 2 | 1.4 |
| Ewing sarcoma | 2 | 1.4 |
| Hepatoblastoma | 2 | 1.4 |
| Non-Hodgkin Lymphoma | 2 | 1.4 |
| Renal Clear cell Carcinoma | 2 | 1.4 |
| Rhabdomyosarcoma (Alveolar) | 2 | 1.4 |
| Abdominal Burkitt Lymphoma | 1 | .7 |
| Acute Lymphocytic Leukemia (T-Cell) | 1 | .7 |
| Anterior mediastinal mass | 1 | .7 |
| Burkett's Lymphoma (Anaplastic lymphoma) | 1 | .7 |
| Burkett's Lymphoma (B-Cell) | 1 | .7 |
| Capillary Haemangioma | 1 | .7 |
| Chondroblastic Osteosarcoma | 1 | .7 |
| Classical Hodgkin Lymphoma (Nodular sclerosing) | 1 | .7 |
| Congenital Neuroblastoma | 1 | .7 |
| Cystic anterior mediastinal mass (Teratoma ) | 1 | .7 |
| Cystic Hygroma | 1 | .7 |
| Dermatofibrosarcoma | 1 | .7 |
| Fibrosarcoma | 1 | .7 |
| Immuture Teratoma | 1 | .7 |
| Infantile Hemangioma | 1 | .7 |
| Lymphoma | 1 | .7 |
| Multiple Endocrine Neoplasia | 1 | .7 |
| Nasopharyngeal carcinoma | 1 | .7 |
| Neuroblastoma. Rhabdomyosarcoma | 1 | .7 |
| Neuroblastoma (Schwanoma ) | 1 | .7 |
| Pineoblastoma | 1 | .7 |
| Rhabdomyosarcoma | 1 | .7 |
| Rhabdomyosarcoma (germ cell tumor ) | 1 | .7 |
| Squamous cell carcinoma | 1 | .7 |
| Thyroid cancer (papillary carcinoma) | 1 | .7 |

**Table 4.5:** Association factors of the disease.

| **History of Exposure** | **Frequency** | **%** |
| --- | --- | --- |
| Missing | 3 | 2.1 |
| No history | 124 | 87.9 |
| Infectious agent: | 14 | 9.9 |
| Malaria | 2 | 1.4 |
| Scabies. Tina capitis | 1 | .7 |
| Tinea corpories | 1 | .7 |
| Tuberculosis | 9 | 6.4 |
| Tuberculosis. Malaria | 1 | .7 |
| **History of chronic illness** |  |  |
| Missing | 2 | 1.4 |
| No | 126 | 89.4 |
| Yes: | 14 | 9.9 |
| Amputation | 1 | .7 |
| Hypertension | 6 | 4.3 |
| Kidney failure | 1 | .7 |
| Sickle cell disease | 4 | 2.8 |
| Thalassemia | 2 | 1.4 |
| **Family history of cancer** |  |  |
| No | 130 | 95.6 |
| yes | 6 | 4.4 |
| Missing | 5 | 3.5 |
| Total | 141 | 100.0 |

**Table 4.6:**
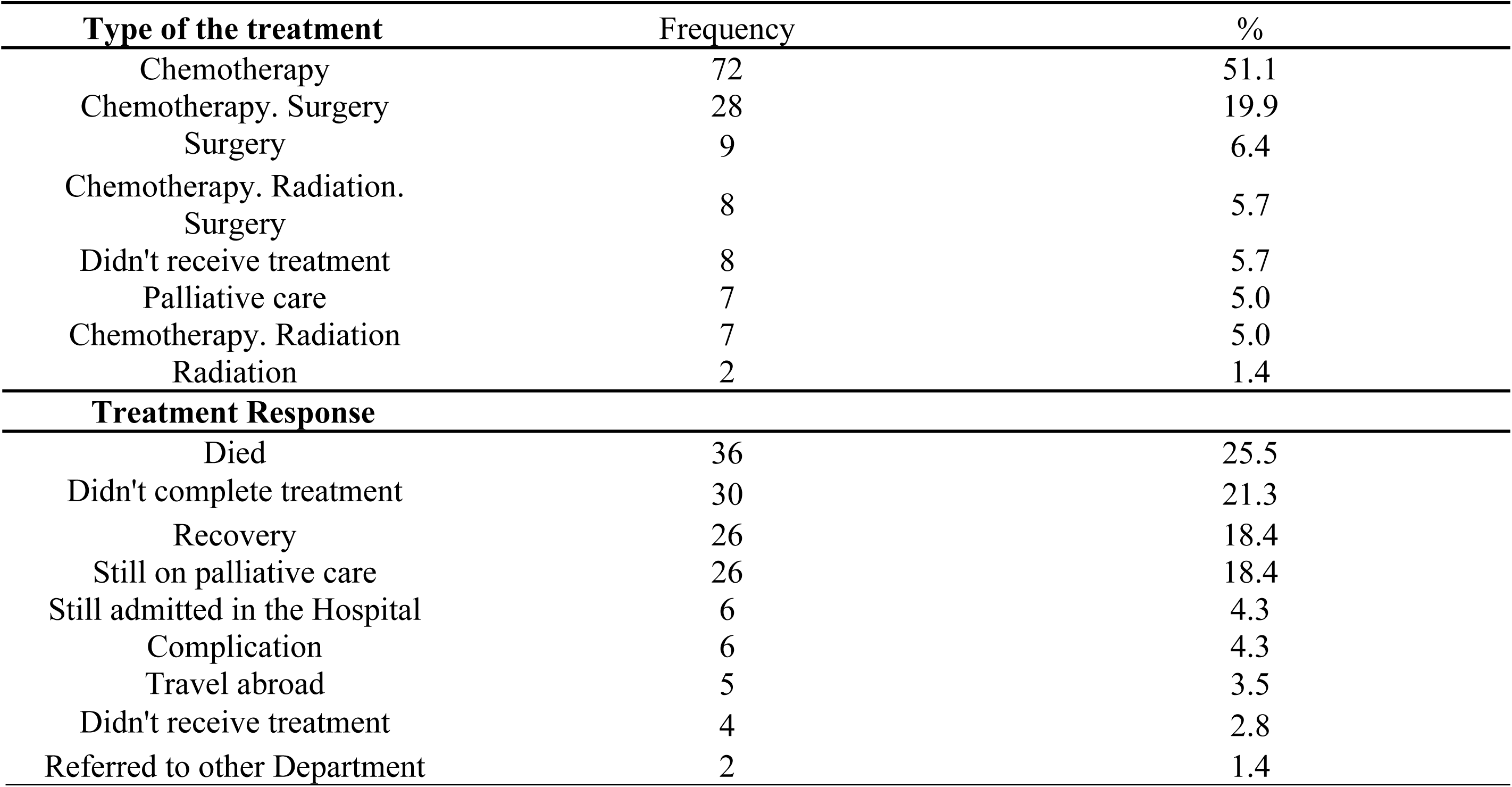
Treatment characteristics.

**Table 4.7:** Cross-tabulation of Gender and the type of cancer.

|  |  | Anterior<br>mediastinal<br>mass | Astrocytoma | Blastoma | Blastoma.<br>sarcoma | Carcinoma | Cystic<br>Hygroma | Hemangioma | Leukemia | Lymphoma | Multiple<br>Endocrine<br>Neoplasia | Sarcoma | Teratoma | Total | p-value |
| --- | --- | --- | --- | --- | --- | --- | --- | --- | --- | --- | --- | --- | --- | --- | --- |
| Male | Count | 1 | 1 | 22 | 0 | 3 | 1 | 0 | 21 | 12 | 1 | 14 | 4 | 80 |  |
|  | % of<br>Total | 0.70% | 0.70% | 15.60% | 0.00% | 2.10% | 0.70% | 0.00% | 14.90% | 8.50% | 0.70% | 9.90% | 2.80% | 56.70% | P-value=<br>0.577 |
| female | Count | 0 | 3 | 19 | 1 | 2 | 0 | 2 | 16 | 7 | 0 | 10 | 1 | 61 |  |
|  | % of<br>Total | 0.00% | 2.10% | 13.50% | 0.70% | 1.40% | 0.00% | 1.40% | 11.30% | 5.00% | 0.00% | 7.10% | 0.70% | 43.30% |  |

**Table 4.8:**
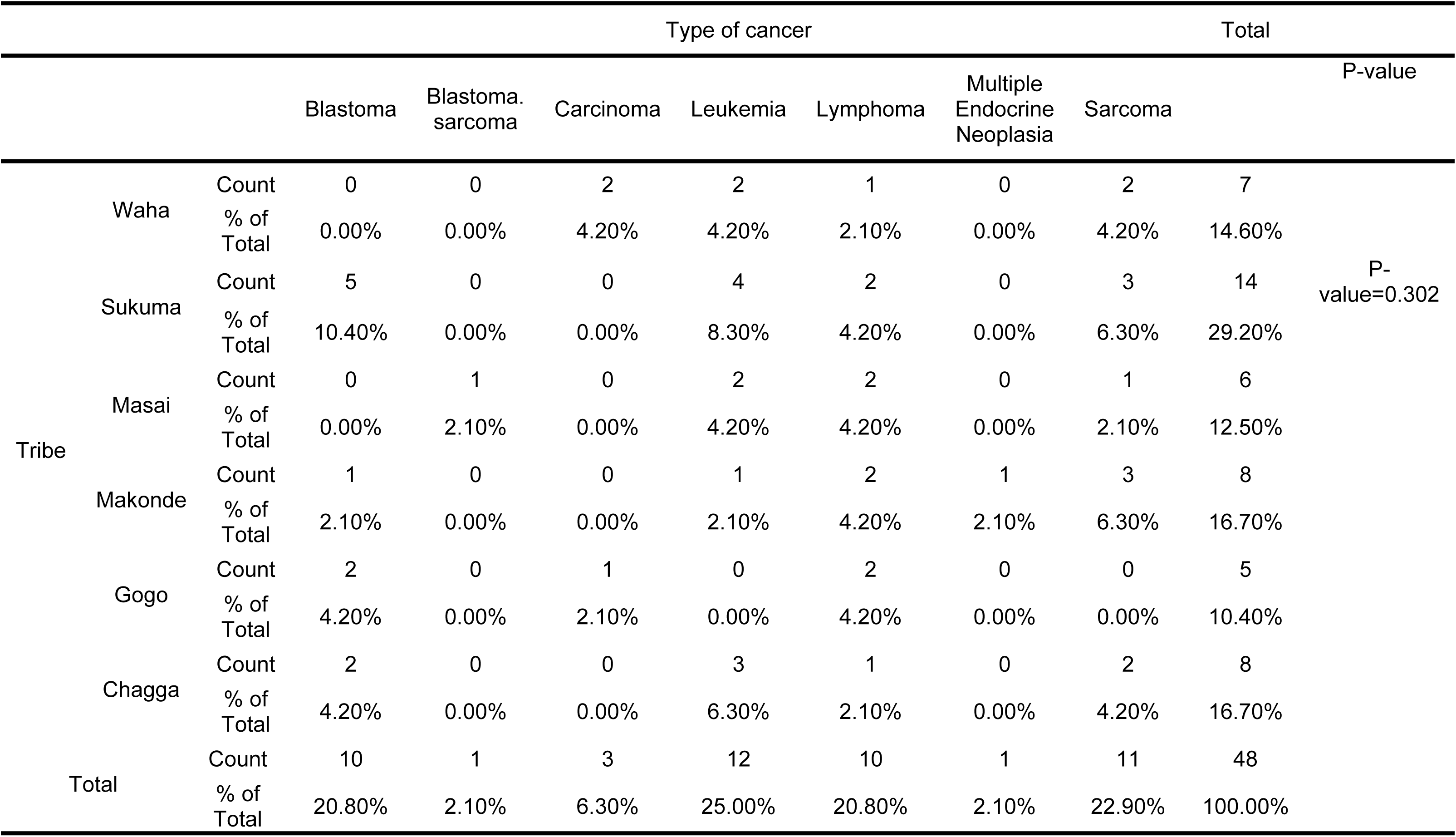
Cross-tabulation of Tribe and Type of cancer.

**Table 4.9:** Cross-tabulation of Admission and the type of cancer.

|  |  | Admission |  |  | Total | p-value |
| --- | --- | --- | --- | --- | --- | --- |
|  |  | 0 | New Admission | Referral |  |  |
| Teratoma | Count | 0 | 1 | 4 | 5 | P-value=0.844 |
|  | % of Total | 0.0% | 0.7% | 2.8% | 3.5% |  |
| Sarcoma | Count | 1 | 7 | 16 | 24 |  |
|  | % of Total | 0.7% | 5.0% | 11.3% | 17.0% |  |
| Multiple Endocrine Neoplasia | Count | 0 | 0 | 1 | 1 |  |
|  | % of Total | 0.0% | 0.0% | 0.7% | 0.7% |  |
| Lymphoma | Count | 0 | 4 | 15 | 19 |  |
|  | % of Total | 0.0% | 2.8% | 10.6% | 13.5% |  |
| Leukemia | Count | 0 | 3 | 34 | 37 |  |
|  | % of Total | 0.0% | 2.1% | 24.1% | 26.2% |  |
| Hemangioma | Count | 0 | 0 | 2 | 2 |  |
|  | % of Total | 0.0% | 0.0% | 1.4% | 1.4% |  |
| Cystic Hygroma | Count | 0 | 1 | 0 | 1 |  |
|  | % of Total | 0.0% | 0.7% | 0.0% | 0.7% |  |
| Carcinoma | Count | 0 | 1 | 4 | 5 |  |
|  | % of Total | 0.0% | 0.7% | 2.8% | 3.5% |  |
| Blastoma.sarcoma | Count | 0 | 0 | 1 | 1 |  |
|  | % of Total | 0.0% | 0.0% | 0.7% | 0.7% |  |
| Blastoma | Count | 0 | 9 | 32 | 41 |  |
|  | % of Total | 0.0% | 6.4% | 22.7% | 29.1% |  |
| Astrocytoma | Count | 0 | 1 | 3 | 4 |  |
|  | % of Total | 0.0% | 0.7% | 2.1% | 2.8% |  |
| Anterior mediastinal mass | Count | 0 | 0 | 1 | 1 |  |
|  | % of Total | 0.0% | 0.0% | 0.7% | 0.7% |  |
| Total | Count | 1 | 27 | 113 | 141 |  |
|  | % of Total | 0.7% | 19.1% | 80.1% | 100.0% |  |

**Table 4.10:** Cross-tabulation of Education Status and Cancer metastasis.

|  |  | Cancer Metastasis |  |  | Total | P-value |
| --- | --- | --- | --- | --- | --- | --- |
|  |  | 0 | No | Yes |  |  |
| 0 | Count | 1 | 22 | 16 | 39 | P-value=0.821 |
|  | % of Total | 0.7% | 15.6% | 11.3% | 27.7% |  |
| college\university | Count | 0 | 9 | 6 | 15 |  |
|  | % of Total | 0.0% | 6.4% | 4.3% | 10.6% |  |
| Illiterate | Count | 0 | 7 | 9 | 16 |  |
|  | % of Total | 0.0% | 5.0% | 6.4% | 11.3% |  |
| Other | Count | 0 | 0 | 1 | 1 |  |
|  | % of Total | 0.0% | 0.0% | 0.7% | 0.7% |  |
| primary school | Count | 1 | 29 | 21 | 51 |  |
|  | % of Total | 0.7% | 20.6% | 14.9% | 36.2% |  |
| secondary school | Count | 0 | 14 | 5 | 19 |  |
|  | % of Total | 0.0% | 9.9% | 3.5% | 13.5% |  |
| Total | Count | 2 | 81 | 58 | 141 |  |
|  | % of Total | 1.4% | 57.4% | 41.1% | 100.0% |  |

**Table 4.11:**
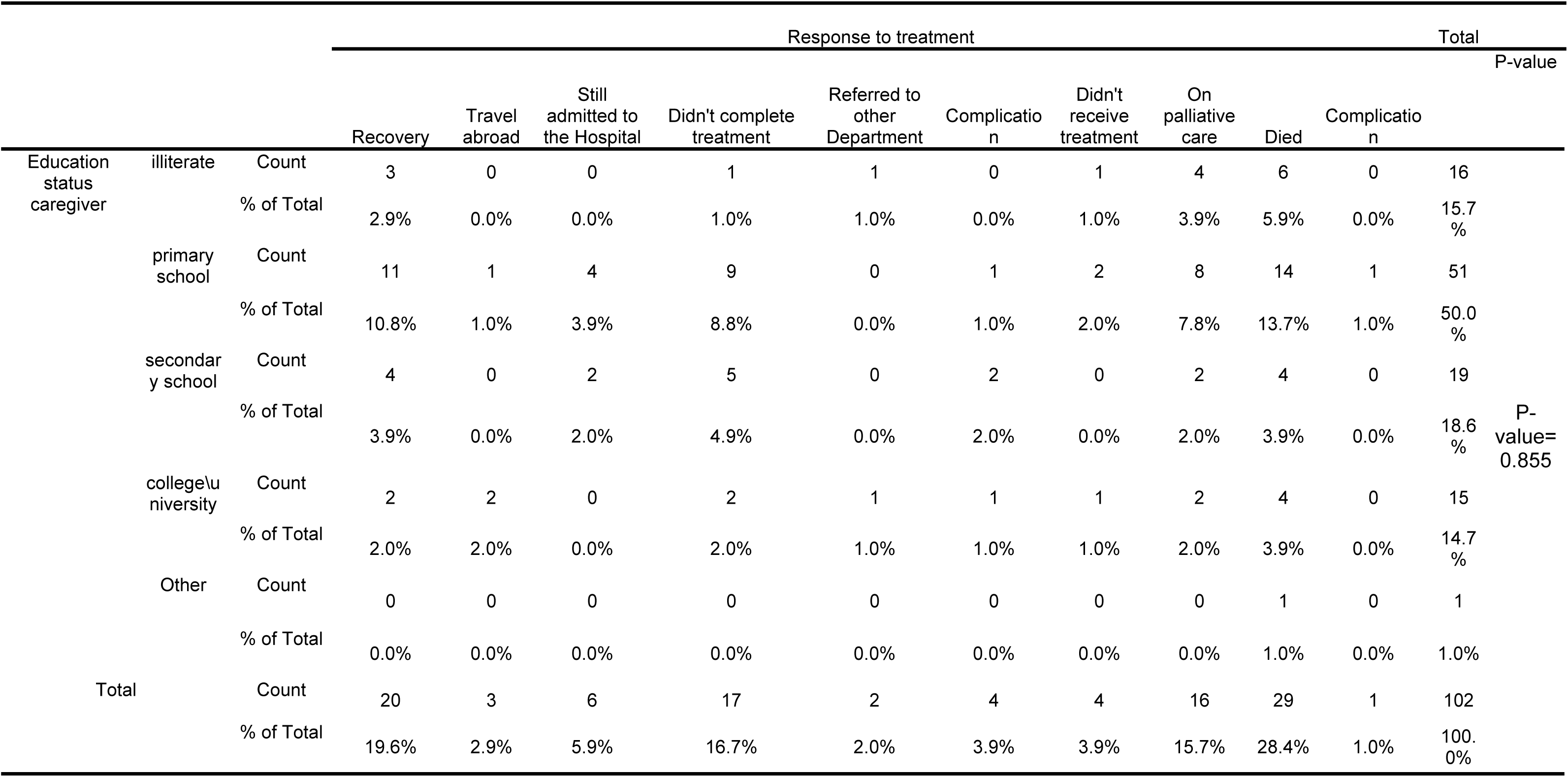
Cross-tabulation of Education status and Response to treatment.

**Table 4.12:**
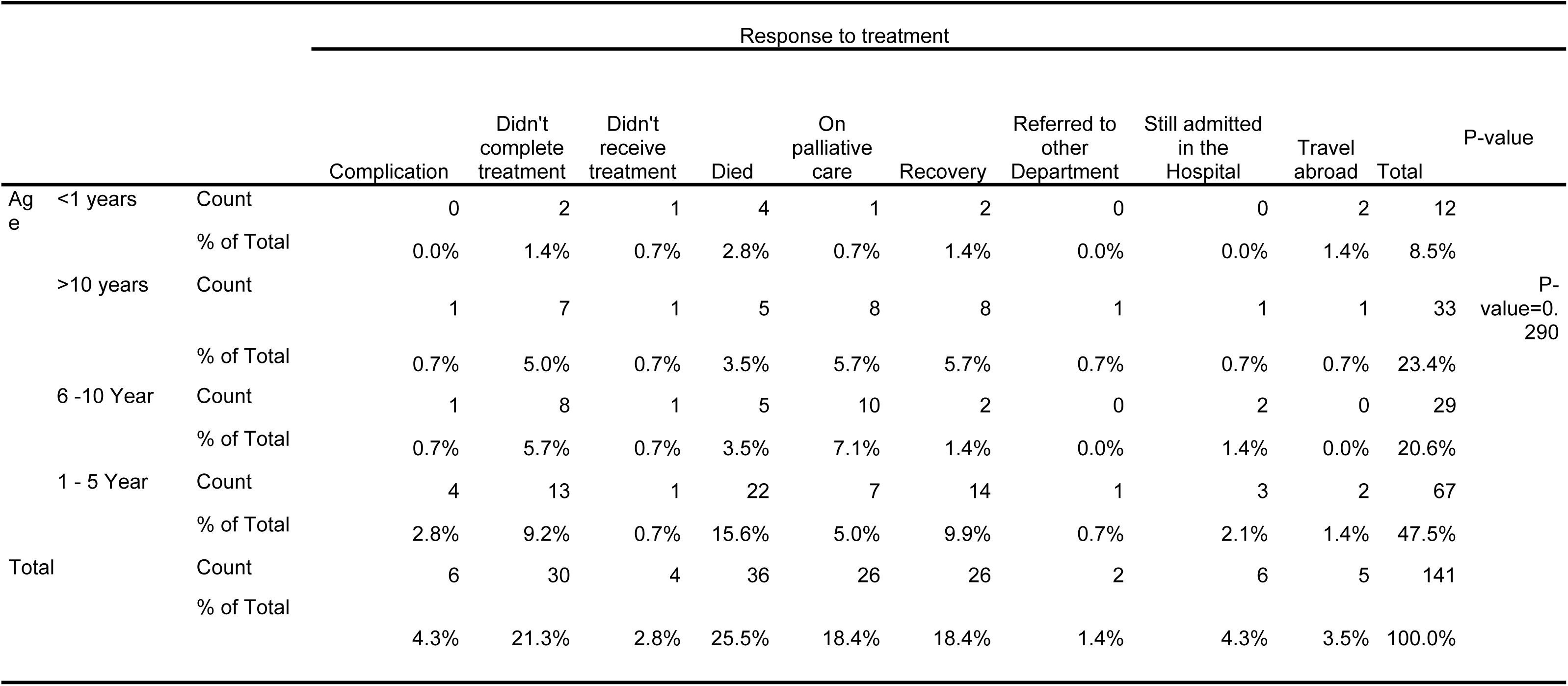
Cross-tabulation of Age and Response to Treatment.

**Table 4.13:**
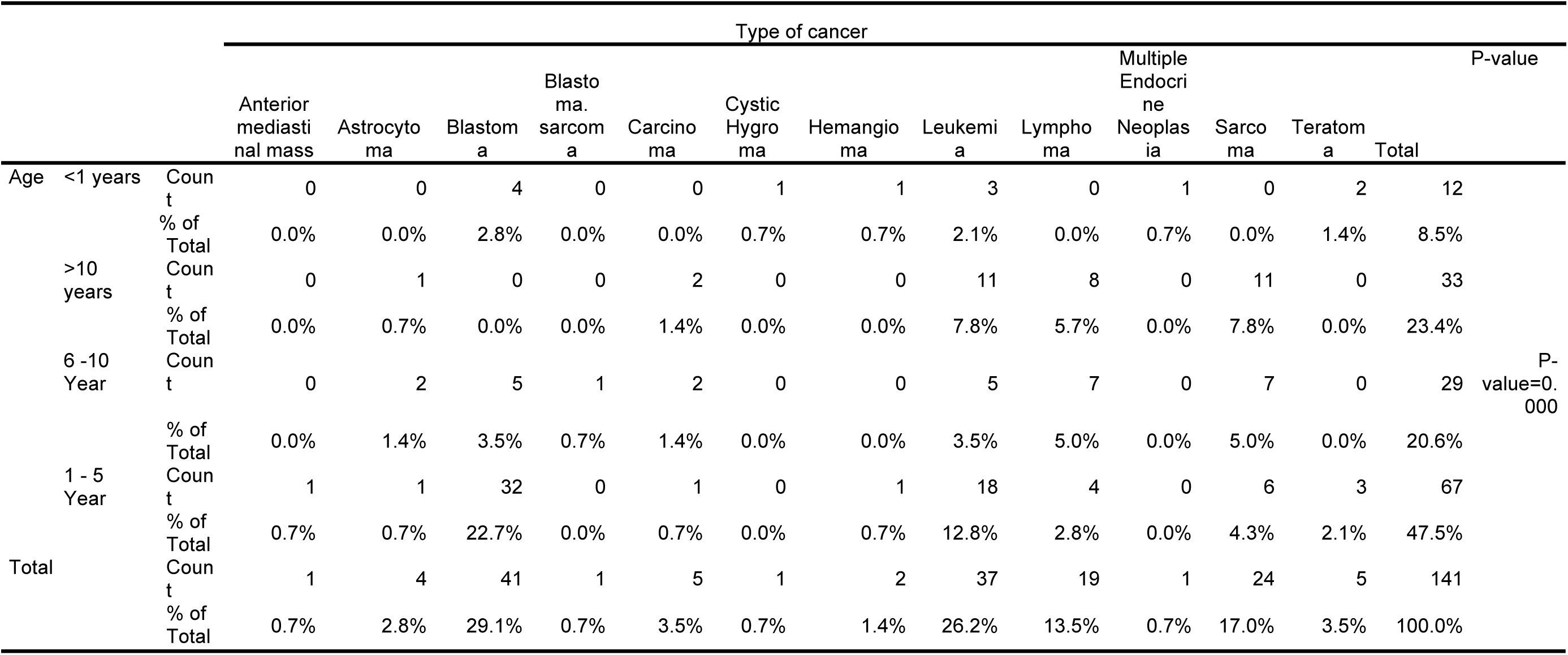
Cross-tabulation of Age and type of cancer.

## DISCUSSION

The current study aimed to describe the demographic characteristics of the patients in the paediatric oncology department of Muhimbili National Hospital, the symptoms that patients presented when they came to the hospital, and the Types of childhood cancer that affected the patients. Thus filling an important gap in this area.

In this study, we found that most of the patients were between 1 and 5 years old, similar to de Oliveira et al.’s study in Brazil, the study in Tanzania, and the study in Bangladesh. There was a male predominance among the patients; this finding is in agreement with several studies. ^6,25,44^ The Sukuma tribe had the highest number of cases, followed by Chagga, Makonde, and Waha. The majority of the patients’ carers get educated until the primary school level, followed by secondary school, which leads to increased cancer mortality because they would not have had the financial means or knowledge to deal with the stress of the illness and its treatment. They came in the late stages. In a cohort study in Italy, the education level of the mother and father was discussed separately, as was their impact on childhood cancer survival. ^41,44^

Dar es Salaam region had the highest number of patient admissions because it is the region where the Muhimbili National Hospital is located, but still, some patients came from other regions like the Pwani (Coast) region (9.2%), Morogoro (6.4%), Dodoma (6.4%), and others. The distance to a cancer treatment facility was inversely linked with the reported regional incidence. Patients with cancer travelled for an average of 4.55 hours to receive care. At the moment, 74.5% of people reside 4 hours away from a cancer care facility, which delays treatment and diagnosis. ^45^

Blastoma has the highest percentage at 29.1%, Leukaemia at 26.2%, and Sarcoma at 17%. A study conducted in Tanzania by Luke Maillie et al. found similar findings: Blastoma and Leukaemia have the highest percentages, and unlike most of the previous research, Leukaemia is the most common type of cancer in children. ^6, 25, 11, 44^, but there were two studies conducted in Tanzania that found that Lymphoma affected a high percentage of patients.^19, 42^ The most common subtypes of Blastoma were Wilm’s tumor (this finding is the same as this article), Retinoblastoma, Leukaemia, Acute Myelogenous Leukaemia, and Acute Lymphocytic Leukaemia (B-cell); and in Sarcoma cancer, the most common subtype was Osteosarcoma. It was a different finding from the article that was done at Muhimbili National Hospital (MNH) in 2019. ^18^ Like most of the previous research, Leukaemia was the most common type of cancer in children. ^6, 25, 11^ male patients have the highest percentage of common cancer types, as the study conducted found that there was no statistically significant difference (p = 0.577) between gender and type of cancer.

The Sukuma tribe has the highest number of patients because it is the biggest tribe in Tanzania. Most of them came with Leukaemia and Blastoma. There was no statistically significant difference (p = 0.302) between tribe and type of cancer.

More than half of the patients were referred from another hospital. 80.1%, Kilimanjaro Christian Medical Centre has the highest number of referral patients because it’s a zonal consultant hospital for the northern region of Tanzania and one of the two cancer centres in northern Tanzania, followed by Mnazi Mmoja Hospital (5.7%). Muhimbili National Hospital is the biggest hospital that has children’s cancer services in Tanzania. There should be centralised treatment in each region because that leads to delays in the diagnosis. There is a study in Japan that discusses this: patients with lengthy trip durations were perceived as incapable of finishing hospital visits and going back home in a single day. Long-distance travel is probably leading to delayed treatment and diagnosis. It is recommended that children and families who must travel a great distance from home should be given special accommodations at neighbouring hospitals, as hospital stays can sometimes last weeks or months. ^33^ There is a shortage of pathologists and paediatric oncologists in Tanzania, which led to referring most of the patients to Muhimbili National Hospital. ^43,19^ Most of the patients were referred for Blastoma (41%), Leukaemia (33.7%), and Sarcoma (22.4%). There was no statistically significant difference (p = 0.577) between patients’ admission and type of cancer.

There was a strong association between patients’ age and type of cancer (p = 0.000). In patients between 1 and 5 years old, most of them had Blastoma (22.7%) and Leukaemia (112.8%), the same result that was found in a study conducted in Brazil. ^28^ According to the World Health Organisation (WHO), the most common types of cancer in children were Leukaemia, Brain cancer, and Lymphoma, ^46^, but in Tanzania, with an incidence of 1.4/100,000 people, the most common childhood cancer was Wilms tumour, Retinoblastoma, and Acute Lymphoplastic Leukaemia,^45^ the same as in this study.

In addition to that, mass is the most common presenting feature (33.8%), followed by abdominal distention (29%), pain (20.9%), and fever (17.3%). Wilms tumour was the most common subtype of Blastoma cancer; in a similar tudy ^39^, most patients presented with abdominal distention (21.9%) or pain (18.8%), and Retinoblastoma was the second most common subtype in Blastoma cancer. Patients presented with a whitish pupillary reflex (18.8%) with fever or eye swelling. Acute Myelogenous Leukaemia is the most common subtype of Leukaemia in patients who present with a fever of 50%; this finding agrees with the study conducted by Sandra Castejon-Ramirez et al.^26^ and generalised body weakness of 35.7%.

A significant proportion of the patients presented with advanced-stage disease, with 50% classified in stage 3 and 32% in stage 4. A study conducted by Thecla W. Kohi et al. A lot of cancer cases in Tanzania are not diagnosed until later because there aren’t many healthcare institutions offering cancer-related care and treatment. ^42^ Research conducted in three populations of sub-Saharan Africa found that the one reason for poor cancer survival in African patients was their diagnosis at an advanced stage. Article conducted by Callum J. R. Mullen et al., The lag of time that effects on the survival and outcome of the study led to a delay in diagnosis, and patients came in late stages. ^30^. Metastasis to other organs was observed in 41% of cases. Notably, the predominant sites of metastasis were the lungs (43.7%) and the central nervous system (19.7%). This advanced stage of presentation may be partially attributed to factors such as the educational background of the carers, with a majority only attaining primary school education, and potentially the socioeconomic challenges prevalent in African contexts. Additionally, the inherent characteristics of paediatric tumours, including rapid growth, short latency periods, and a propensity for invasiveness, may contribute to reduced time for differential diagnosis. ^30^ Statistical analysis revealed no significant association between the presence of metastasis, the stage of cancer, and the education level of the carer (p = 0.821, p = 0.818, respectively). This lack of correlation could be influenced by incomplete data regarding the caregivers’ educational status and the stages of cancer in the recorded data.

Most of the patients didn’t have a history of chronic illness, a family history of cancer, or a history of exposure. Hypertension (4.3%) and sickle cell disease (2.8%) were the most common chronic diseases among patients. Only exposure to infectious agents like tuberculosis (6.4%) was found in our study. There were no findings of other types of exposure, like radiation, chemicals, pesticides, or others. Several studies discuss different types of exposures, like infectious and lifestyle exposure ^17^, pesticide exposure ^2^, phototherapy exposure in pregnancy ^15^, environmental and hereditary influences ^29^, and HIV exposure ^20^. The history of exposure is important, especially in cancer cases, to exclude the cause. There was no statistically significant difference between a history of chronic illness and a history of exposure to a type of cancer (p = 0.466 and p = 0.394), respectively. There was an association and statistically significant difference (p = 0.004) between family history and type of cancer. In Sweden, a study conducted on childhood leukaemia patients with a family history had the same inding. ^38^

Almost half of the patients received chemotherapy (51.1%), the same as the study conducted by Olívia Lopes et al, ^32^ chemotherapy with surgery (19.9%), or surgery alone. However, the majority of the patients died at a rate of 25.5%. Several studies agree on this point: ^13, 23, 24, 34,35^ patients did not complete treatment for many reasons; ^8, 31, 23^ patients didn’t complete treatment due to financial problems. There are studies explaining that ^10, 12, 19^, or waiting for the next session of treatment, recovery patients, and palliative care patients had equal proportions (18.4%).

In this study, it was observed that the cohort comprising patients aged 1 to 5 years exhibited the highest mortality rate at 17.1%, alongside a notable recovery rate of 12.4%. Predominantly, fatalities were recorded in cases where the carer’s educational attainment was limited to primary school, potentially indicative of impediments to treatment efficacy. Furthermore, statistical analysis revealed no significant correlation between the treatment outcomes and variables such as the carer’s educational status or the patient’s age, with p-values of 0.855 and 0.290, respectively. There was an association (p = 0.006) between the cancer stage and response to treatment because most of them came in late-stage disease with metastasis cancer; these two studies agreed with that ^20, 36^ and some of the patients died before receiving treatment due to financial problems. Several factors, including high prices, a lack of health insurance, and restricted financial resources, make the detection and treatment of children’s cancer in low- and middle-income nations difficult. Out-of-pocket medical costs are a source of stress for families and have a detrimental impact on the quality of life and treatment outcomes for cancer patients. The long-term impact of these costs on the family’s financial security. ^37^

The limitation of this study was due to the exclusion of other types of tumours in the paediatric department of the Muhimbili National Hospital (MNH) and the fact that it didn’t complete all the record data in the study period because there were missing files and a lot of information missing in the record data. Even after a call to the family of the patients, most of them didn’t respond or didn’t know. To assess the success of the research, there should be full coverage data for at least 1 year, but due to the limitation of graduation time, the research was conducted in 4 months only.

This study can inform clinical practice and policy changes in several significant ways. The identification of common cancer types and their clinical presentations among children can aid in refining diagnostic protocols, leading to earlier and more accurate diagnoses. Moreover, understanding the prevalent cancer types and their responses to various treatments allows for the development of more effective, tailored treatment strategies. Our findings can guide the allocation of resources, such as diagnostic radiology and staff training, to areas where they are most needed. We believe that data on demographic characteristics and associated factors can inform public health policies, focusing on prevention and screening strategies and early detection programs. Insights from our study can direct future research, addressing specific challenges identified in the treatment and management of childhood cancers at the MNH.

## CONCLUSION

Childhood cancer in Tanzania presents with high rates of mortality and morbidity. The study at Muhimbili National Hospital highlights that many patients are diagnosed at a late stage, which significantly reduces their chances of survival. The predominant cancers identified were Blastoma and Leukemia, primarily affecting children aged 1 to 5 years. This delayed diagnosis is often compounded by socioeconomic challenges such as limited financial resources and low educational levels among caregivers, which hinder the completion of treatment.

Additionally, the research emphasizes the lack of comprehensive medical records and the need for improved data management to ensure accurate and complete patient histories. Enhancing early detection methods and providing adequate financial and educational support for families could improve treatment outcomes. Implementing public health strategies focused on early cancer detection, better diagnostic facilities, and trained medical personnel are crucial steps towards reducing the burden of childhood cancer in Tanzania.

The findings underscore the necessity of addressing both medical and socioeconomic barriers to improve the overall healthcare system and patient outcomes in pediatric oncology. Future research should focus on long-term data collection to provide a more detailed understanding of childhood cancer trends and the effectiveness of interventions.

## RECOMMENDATIONS

We recommend evaluating the record data system to ensure that no patient files are missing. Getting a good history from the patient when he came to the hospital helped in excluding the cause of the disease and informed the patient’s family about the disease. Review the type of treatment to see if there is any drug resistance or financial support for poor families that will reduce the high percentage of patients dying and improve the morbidity and mortality of the disease in children.

The findings suggest an urgent need for enhanced public health strategies focusing on early cancer detection and education about pediatric cancers. The study underscores the necessity of improved diagnostic facilities and trained medical personnel within Tanzania to facilitate early and accurate cancer diagnosis. Additionally, public health policies must address the socioeconomic barriers that contribute to late-stage cancer presentation in children.

UMST University of Medical Sciences and Technology

MNH Muhimbili National Hospital

14C The international childhood cancer cohort consortium

LMICs Low and medial income countries

## Data Availability

All relevant data are within the manuscript and its Supporting Information files

## DECLARATIONS

### Ethics Approval and Consent to participate

The approval from the hospital administration has been granted by the medical director’s office at Muhimbili National Hospital (MNH). As well as the approval of the research department. The proposal was then submitted to the research office at the Ministry of Health, Dar es Salaam State, to get ethical approval for the study.

All patients’ data will be collected anonymously through a data collection tool. Due to the cross-sectional study design, there were patient files. The participants’ confidentiality was assured with the use of an anonymous data collection tool called Codes to protect the anonymity of the patients.

### Consent for publication

All authors have read the final manuscript and given their consent for this article to be published in this journal. No clinical details of participants that might compromise their anonymity were used in the development of this manuscript titled **“Presentation and Types of childhood cancer at The Muhimbili National Hospital (MNH), 2023”.**

### Availability of supporting data

All supporting data are available.

### Authors’ contributions

All authors have read the final manuscript and given their approval for publication.

## ACKNOWLEDGMENTS

I would like to express my special thanks and gratitude to my supervisor, Dr. Yasar Hammor, who made this work possible and provided invaluable guidance throughout this research. I would also like to thank Dr. Rehema H. Laiti and my co-supervisors, Dr. Gahada and Prof. Amin Alagib Mohammed. I would also like to give my warmest thanks to my youngest brother, who was one of the pediatric cancer patients and inspired me to do this research.

---

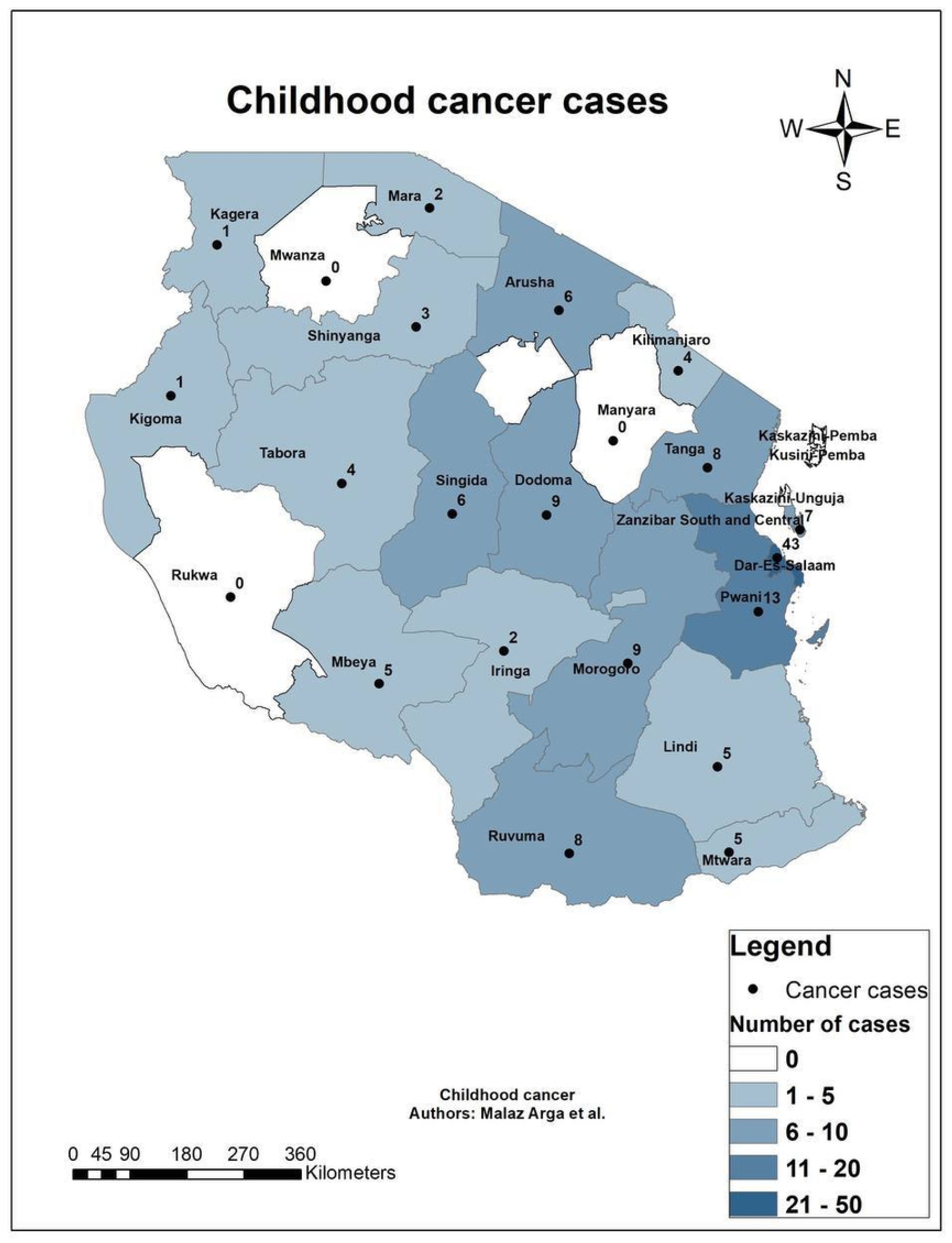

